# Vaccine effectiveness against hospital admission in South African health care workers who received a homologous booster of Ad26.COV2 during an Omicron COVID19 wave: Preliminary Results of the Sisonke 2 Study

**DOI:** 10.1101/2021.12.28.21268436

**Authors:** Glenda E Gray, Shirley Collie, Nigel Garrett, Ameena Goga, Jared Champion, Matthew Zylstra, Tarylee Reddy, Nonhlanhla Yende, Ishen Seocharan, Azwi Takalani, Ian Sanne, Fatima Mayat, Jacky Odhiambo, Lesley Bamford, Harry Moultrie, Lara Fairall, Linda-Gail Bekker, on behalf of the Sisonke Study Team

## Abstract

Following the results of the ENSEMBLE 2 study, which demonstrated improved vaccine efficacy of a two-dose regimen of Ad26.COV.2 vaccine given 2 months apart, we expanded the Sisonke study which had provided single dose Ad26.COV.2 vaccine to almost 500 000 health care workers (HCW) in South Africa to include a booster dose of the Ad26.COV.2. Sisonke 2 enrolled 227 310 HCW from the 8 November to the 17 December 2021. Enrolment commenced before the onset of the Omicron driven fourth wave in South Africa affording us an opportunity to evaluate early VE in preventing hospital admissions of a homologous boost of the Ad26.COV.2 vaccine given 6-9 months after the initial vaccination in HCW.

We estimated vaccine effectiveness (VE) of the Ad26.COV2.S vaccine booster in 69 092 HCW as compared to unvaccinated individuals enrolled in the same managed care organization using a test negative design. We compared VE against COVID19 admission for omicron during the period 15 November to 20 December 2021. After adjusting for confounders, we observed that VE for hospitalisation increased over time since booster dose, from 63% (95%CI 31-81%); to 84% (95% CI 67-92%) and then 85% (95% CI: 54-95%), 0-13 days, 14-27 days, and 1-2 months post-boost.

We provide the first evidence of the effectiveness of a homologous Ad26.COV.2 vaccine boost given 6-9 months after the initial single vaccination series during a period of omicron variant circulation. This data is important given the increased reliance on the Ad26.COV.2 vaccine in Africa.

## Commentary

South Africa is in the middle of a fourth wave of the COVID19 pandemic, driven by Omicron, a new variant of concern that emerged in early November 2021, and has spread rapidly across the world.^1-5^ Both in-vitro experiments and epidemiological data suggest that Omicron may reduce vaccine effectiveness (VE).^6-8^ When the South African national vaccine roll-out faltered in February 2021, we were able to vaccinate health care workers (HCW), in a phase 3B open-label implementation trial, utilizing a single-dose of the Ad26.COV.2 vaccine. This study, known as the Sisonke Study, based on efficacy data from the ENSEMBLE study^9^, enrolled 477 234 HCWs from the 17 February to the 17 May 2021, established vaccine effectiveness against both Delta and Beta^10^. Following the results of the ENSEMBLE 2^11^ study, which demonstrated improved vaccine efficacy of a two-dose regimen of Ad26.COV.2 vaccine given 2 months apart, we expanded the Sisonke study (Sisonke 2) to include a booster dose of the Ad26.COV.2. Sisonke 2 enrolled 227 310 HCW who had received an initial single dose of the Ad26.COV2. S vaccine, from the 8 November to the 17 December 2021. Enrolment commenced before the onset of the Omicron driven fourth wave in South Africa affording us an opportunity to evaluate early VE in preventing hospital admissions of a homologous boost of the Ad26.COV.2 vaccine given 6-9 months after the initial vaccination in HCW.

Sisonke 2 was conducted in approximately 350 vaccination centres, supported by 43 clinical research sites, across all nine provinces of South Africa. The study was approved by all pertinent regulatory and ethical review committees. Utilising data from Discovery Health, a South African managed care organization, we estimated vaccine effectiveness (VE) of the Ad26.COV2.S vaccine booster in 69 092 HCW as compared to unvaccinated individuals enrolled in the same managed care organization. We compared VE against COVID19 admission for omicron during the period 15 November to 20 December 2021. (Table 1) Datasets utilized included COVID19 PCR test results, pre-authorization admission data, full history of members claims records, chronic disease registrations, and body mass index data to obtain the number of COVID19 risk factors per individual.

**Table 1:** Study population showing test positivity and hospital admissions for COVID-19 during the Omicron period.

| (n, %) | Number of tests | Number of positive COVID19 tests | Number of COVID19 admissions |
| --- | --- | --- | --- |
| **Total** | 52,468 (100.0) | 20,021 (100.0) | 734 (100.0) |
| **Vaccination status** |  |  |  |
| Not Vaccinated | 47,332 (90.2) | 18,294 (91.4) | 713 (97.1) |
| J&J - 0 to 13 days post dose 2 | 1,799 (3.4) | 540 (2.7) | 10 (1.4) |
| J&J - 14 to 27 days post dose 2 | 2,514 (4.8) | 881 (4.4) | 8 (1.1) |
| J&J -1-2 months (27 to 87 days) post dose 2 | 823 (1.6) | 306 (1.5) | 3 (0.4) |
| **Age (in years)** |  |  |  |
| 18-29 | 13,278 (25.3) | 5,605 (28.0) | 121 (16.5) |
| 30-39 | 18,899 (36.0) | 7,649 (38.2) | 185 (25.2) |
| 40-49 | 9,806 (18.7) | 3,847 (19.2) | 128 (17.4) |
| 50-59 | 5,339 (10.2) | 1,750 (8.7) | 95 (12.9) |
| 60-69 | 2,827 (5.4) | 703 (3.5) | 76 (10.4) |
| 70-79 | 1,582 (3.0) | 323 (1.6) | 76 (10.4) |
| 80+ | 737 (1.4) | 144 (0.7) | 53 (7.2) |
| **Sex** |  |  |  |
| Male | 20,585 (39.2) | 8,430 (42.1) | 304 (41.4) |
| Female | 31,883 (60.8) | 11,591 (57.9) | 430 (58.6) |
| **Number of CDC risk factors\*\*** |  |  |  |
| 0 | 36,467 (69.5) | 14,768 (73.8) | 362 (49.3) |
| 1 | 11,396 (21.7) | 4,026 (20.1) | 211 (28.7) |
| 2 | 3,375 (6.4) | 984 (4.9) | 107 (14.6) |
| 3+ | 1,230 (2.3) | 243 (1.2) | 54 (7.4) |
| **Calendar week** |  |  |  |
| 46 | 4,512 (8.6) | 165 (0.8) | 11 (1.5) |
| 47 | 5,681 (10.8) | 887 (4.4) | 32 (4.4) |
| 48 | 11,552 (22.0) | 4,514 (22.5) | 145 (19.8) |
| 49 | 16,820 (32.1) | 7,741 (38.7) | 257 (35.0) |
| 50 | 11,759 (22.4) | 5,636 (28.2) | 241 (32.8) |
| 51 | 2,144 (4.1) | 1,078 (5.4) | 48 (6.5) |
| **Period of last documented prior infection** |  |  |  |
| None | 43,179 (82.3) | 17,274 (86.3) | 664 (90.5) |
| Ancestral (2020/03/01-2020/10/31) | 2,088 (4.0) | 812 (4.1) | 24 (3.3) |
| Beta (2020/11/01-2021/05/16) | 2,483 (4.7) | 785 (3.9) | 24 (3.3) |
| Delta (2021/05/17-2021/10/31) | 4,718 (9.0) | 1,150 (5.7) | 22 (3.0) |
| **Province** |  |  |  |
| Eastern Cape | 2,252 (4.3) | 823 (4.1) | 22 (3.0) |
| Free State | 2,053 (3.9) | 691 (3.5) | 37 (5.0) |
| Gauteng | 25,154 (47.9) | 10,382 (51.9) | 325 (44.3) |
| KwaZulu Natal | 7,575 (14.4) | 2,521 (12.6) | 169 (23.0) |
| Limpopo | 1,364 (2.6) | 643 (3.2) | 18 (2.5) |
| Mpumalanga | 2,563 (4.9) | 1,077 (5.4) | 38 (5.2) |
| Northwest | 2,147 (4.1) | 847 (4.2) | 41 (5.6) |
| Northern Cape | 875 (1.7) | 242 (1.2) | 6 (0.8) |
| Western Cape | 7,904 (15.1) | 2,507 (12.5) | 74 (10.1) |
| Not allocated | 581 (1.1) | 288 (1.4) | 4 (0.5) |

*\*\*see supplementary material for clinical definition of number of CDC risk factors*

We applied a test-negative design and data exclusion rules to obtain VE estimates^12^. Vaccine effectiveness estimates were obtained from the following formula: 1-odds of COVID19 admission amongst the vaccinated population, where the odds ratio was calculated using logistic regression, and adjusting for confounders of age (18-29, then 10-year age bands and then age 80+), sex, number of documented CDC risk factors (0,1,2,3+), surveillance week, period of prior documented infection (D614G:2020/03/01-2020/10/31, Beta:2020/11/01-2021/05/16, and Delta: 2021/05/17-2021/10/31), and geographic region (province). COVID19 related admission was a dependent variable, while vaccination status was included as an independent variable.

Vaccination status was analyzed in the following categories: not vaccinated; 0-6, 7-13, 14-27 days, and 1 to 2 months since second dose vaccination for Ad26.COV2 recipients. Single dose and other vaccine recipients, and those vaccinated in the public sector based on a match with the national Electronic Vaccination Data System (EVDS) up to 25 August 2021, were excluded from the analysis. Unvaccinated controls may have included vaccinated individuals since 25 August 2021.

We excluded anyone with a negative COVID19 tests within 21 days of a positive test; or negative tests within 7 days of another negative test result; or positive and negative results within 90 days of a previous positive test result in the analysis. For any individuals with more than 3 test results a random selection of three of those results were included. Finally, indeterminate test results, and test results for individuals younger than age 18 years were also excluded. We performed a sensitivity analysis using only COVID19 PCR results within the Gauteng province, given the geographical concentration of the Omicron variant in that province over the study period. Calculations were performed using R version 4.1.1

After adjusting for confounders, we observed that VE for hospitalisation increased over time since booster dose, from 63% (95%CI 31-81%); to 84% (95% CI 67-92%) and then 85% (95% CI: 54-95%), 0-13 days, 14-27 days, and 1-2 months post-boost (Table 2). Preliminary data emanating from pseudovirus neutralization assays have shown the ability of Omicron to escape antibody neutralization after a single dose Ad26.COV.2 vaccine (Penny Moore, personal communication), thereby raising concerns about the impact that this variant will have on VE. Our data demonstrates that a homologous boost given to HCW 6-9 months after the initial Ad26.COV.2 vaccine is protective against hospital admissions.

**Table 2:**
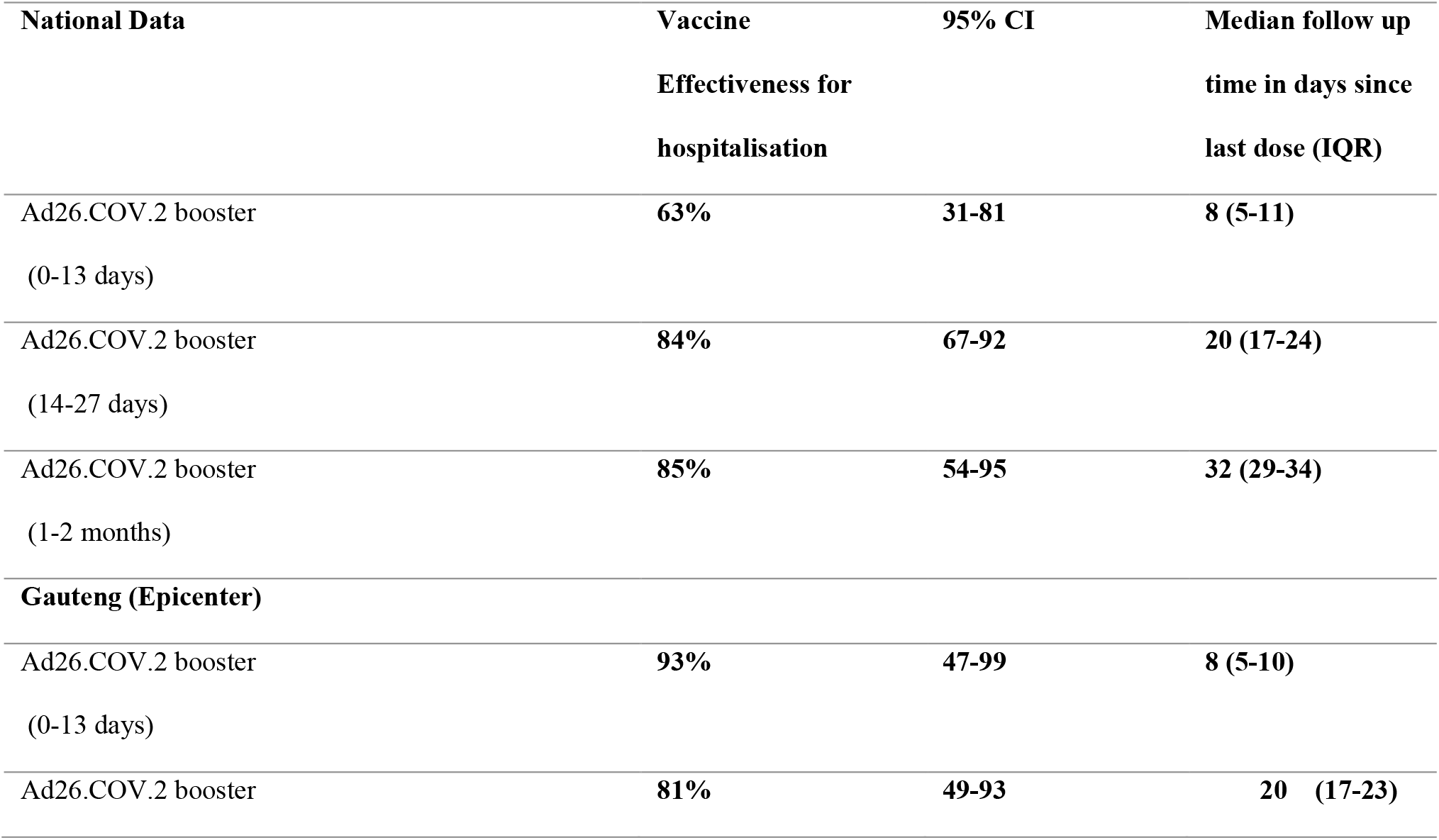
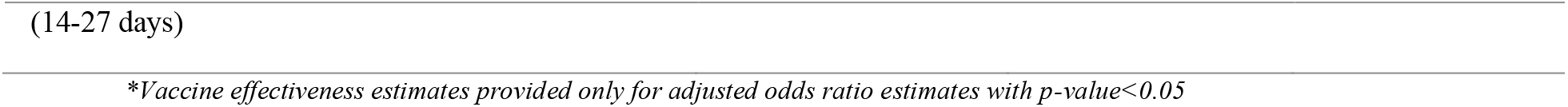
National and Gauteng (Epicenter) Vaccine Effectiveness against hospitalization of homologous Ad26.COV.2 booster over time.

| **National Data** | **Vaccine Effectiveness for hospitalisation** | **95% CI** | **Median follow up time in days since last dose (IQR)** |
| --- | --- | --- | --- |
| Ad26.COV.2 booster<br>(0-13 days) | **63%** | **31-81** | **8 (5-11)** |
| Ad26.COV.2 booster<br>(14-27 days) | **84%** | **67-92** | **20 (17-24)** |
| Ad26.COV.2 booster<br>(1-2 months) | **85%** | **54-95** | **32 (29-34)** |
| **Gauteng (Epicenter)** |  |  |  |
| Ad26.COV.2 booster<br>(0-13 days) | **93%** | **47-99** | **8 (5-10)** |
| Ad26.COV.2 booster | **81%** | **49-93** | **20 (17-23)** |

Our analysis has several limitations: overall follow up was short for Ad26.COV.2 vaccine boosts-median follow up since the boost was 8 days (IQR 5-11) for HCW (n=1 799) who received their boost within 0-13 days; 20 days (IQR 17-24) for those 14-27 days post boost (n= 2 514) and 32 days (IQR 29-34) for those who had been boosted for 1-2 months (n=823) which could impact overall VE. Omicron dominance was also increasing around the country over this time but started from a low Delta variant base in Gauteng and quickly became the dominant variant in that province and around the country, and as such similar results in Gauteng are reassuring; in addition, as we did not have access to the national EVDS some of the control individuals after late August may have been vaccinated leading to a potential underestimation of VE.

We provide the first evidence of the effectiveness of a homologous Ad26.COV.2 vaccine boost given 6-9 months after the initial single vaccination series during a period of omicron variant circulation. We have demonstrated that VE for hospital admissions increased over time up to at least 1-2 months post-boost. This increase in VE is consistent with other studies that have evaluated the Ad26COV.2 vaccine, showing consistency in protection against severe disease over time, indicative that protection against severe disease may be due to cellular immunity and immune memory rather than neutralizing antibodies^9,11,12^. This data is important given the increased reliance on the Ad26.COV.2 vaccine in Africa.

## Data Availability

All data produced in the present study are available upon reasonable request to the authors

## Funding and Analyses

Funding was provided by the National Treasury of South Africa, the National Department of Health, Solidarity Response Fund NPC, The Michael & Susan Dell Foundation, The Elma Vaccines and Immunization Foundation - Grant number 21-V0001, and the Bill & Melinda Gates Foundation – grant number INV-030342. The content is solely the responsibility of the authors and does not necessarily represent the official views of the Johnson and Johnson or our funders. The funders had an opportunity to review a preliminary version of the manuscript, the authors are solely responsible for the final content and interpretation. Data analyses were performed by SC, JC and MZ. The manuscript first draft was written by GG, SC and LGB and all authors reviewed and gave input to the subsequent drafts. All authors signed off on the final copy. No competing interests with regard to this study have been declared.

## Acknowledgements

The authors wish to thank the Health Care Workers who participated in the Sisonke Study. Appreciation to the clinical research site investigators, the study staff and teams and the support staff at the SAMRC. We are deeply grateful for the assistance of Paul Stoffels, Johan van Hoof and Abeda Williams from Janssen, Johnson and Johnson who facilitated and provided the investigational product. We also wish to acknowledge our regulator, SAHPRA as well as the Health Research Ethics Committees who provided guidance and oversight.

## Supplement

**Supplementary table 1.** **The following list of comorbidities (presented in alphabetical order) has been adapted from the Prescribed Minimum Benefits Chronic Disease List (CDL) and the Centers for Disease Control and Prevention (CDC) list of conditions associated with increased risk of severe COVID-19 (https://www.cdc.gov/coronavirus/2019-ncov/need-extra-precautions/people-with-medical-conditions.html).

| Number | Category | Conditions |
| --- | --- | --- |
| 1 | Cancer | • Cancer |
| 2 | Cardiovascular disease | • Cardiac failure<br>• Cardiomyopathy<br>• Coronary artery disease<br>• Dysrhythmias<br>• Peripheral arterial disease<br>• Cerebrovascular disease (including stroke) |
| 3 | Chronic renal disease | • Chronic renal disease |
| 4 | Chronic respiratory disease | • Asthma<br>• COPD<br>• Bronchiectasis |
| 5 | Diabetes mellitus | • Diabetes Mellitus 1<br>• Diabetes Mellitus 2 |
| 6 | HIV | • HIV |
| 7 | Hypertension | • Hypertension |
| 8 | Liver disease | • Alcoholic liver disease<br>• Fatty liver disease<br>• Cirrhosis |
| 9 | Neurological disorders | • Epilepsy<br>• Parkinson's disease<br>• Dementia (any cause, including Alzheimer's disease) |
| 10 | Overweight / obesity | • BMI >25 |
| 11 | Severe mental disorders | • Bipolar mood disorder<br>• Schizophrenia |
| 12 | Solid organ transplant | • History of Kidney, liver, heart, or lung transplant |

**Supplementary Table 2:**
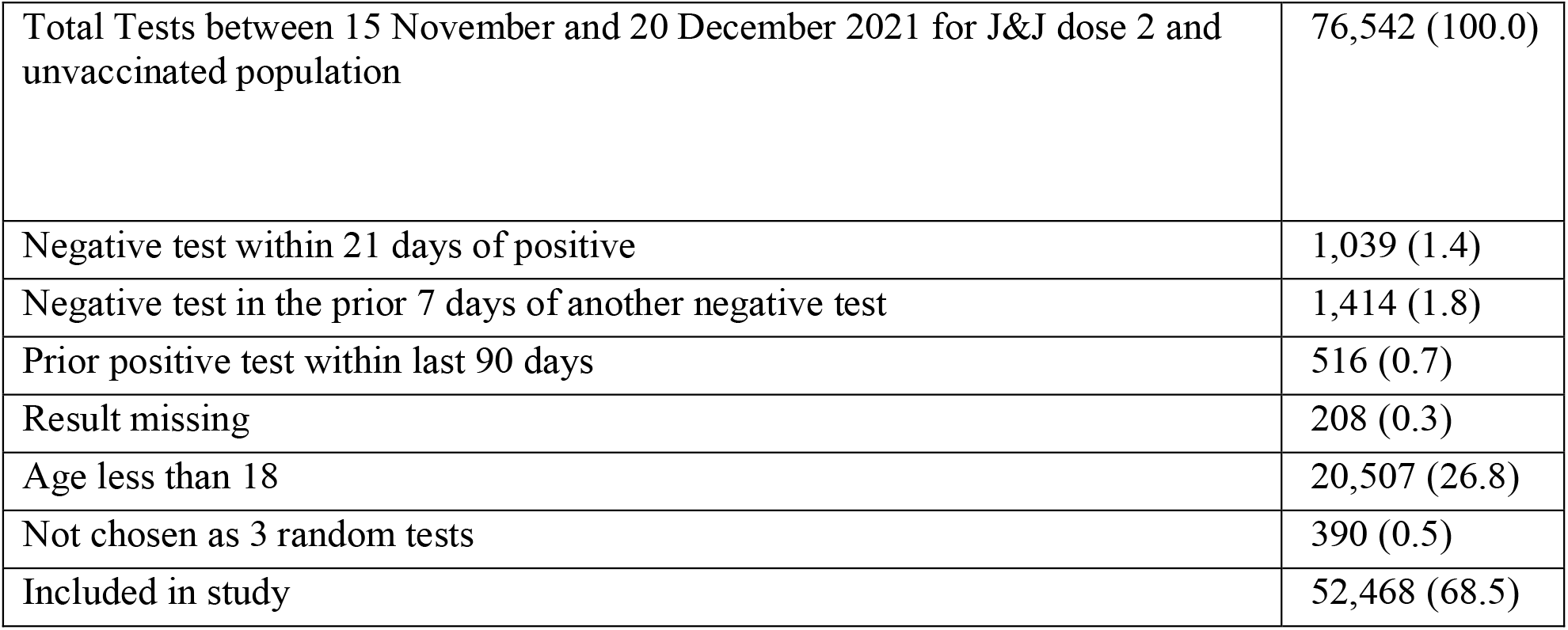
Exclusion volumes.

**Supplementary Table 3:** Volumes of sensitivity for Gauteng:

|  | Number of tests | Number of positive COVID19 tests | Number of COVID19 admissions |
| --- | --- | --- | --- |
| Not Vaccinated | 22,903 (91.1) | 9,652 (93.0) | 320 (98.5) |
| J&J - 0 to 13 days post dose 2 | 882 (3.5) | 269 (2.6) | 1 (0.3) |
| J&J - 14 to 27 days post dose 2 | 1,099 (4.4) | 375 (3.6) | 4 (1.2) |
| J&J -27 to 87 post dose 2 | 270 (1.1) | 86 (0.8) | 0 (0.0) |
| Total | 25,154 (100.0) | 10,382 (100.0) | 325 (100.0) |

## References

1. Chotiner I. How South African Researchers Identified the Omicron Variant of COVID. The New Yorker. 2021 30 November 2021. Available from https://www.newyorker.com/news/q-and-a/how-south-african-researchers-identified-the-omicron-variant-of-covid. Accessed 17 December 2021.

2. Callaway E. Heavily mutated Omicron variant puts scientists on alert. Nature Magazine. 2021. Available from https://www.scientificamerican.com/article/heavily-mutated-omicron-variant-puts-scientists-on-alert/. Accessed 17 December 2021.

3. World Health Organization. Classification of Omicron (B.1.1.529): SARS-CoV-2 Variant of Concern. Available from https://www.who.int/news/item/26-11-2021-classification-of-omicron-(b.1.1.529)-sars-cov-2-variant-of-concern. Accesed 17 December 2021.

4. Grabowski F, Kochańczyk M, Lipniacki T. Omicron strain spreads with the doubling time of 3.2—3.6 days in South Africa province of Gauteng that achieved herd immunity to Delta variant. MedRxiv 2021: doi: https://doi.org/10.1101/2021.12.08.21267494.

5. de Oliveira T, Venter M, Bhiman J, Scheepers C, Preiser W. Here’s what Omicron can tell us about how COVID-19 variants are discovered. World Economic Forum, 2021. Available from https://www.weforum.org/agenda/2021/11/coronavirus-variant-discovery-omicron-health/. Accessed 17 December 2021.

6. Callaway E. Omicron likely to weaken COVID vaccine protection. Nature. 2021. Available from https://www.nature.com/articles/d41586-021-03672-3. Accessed 17 December 2021

7. Wilhelm A, Widera M, Grikscheit K, et al. Reduced Neutralization of SARS-CoV-2 Omicron Variant by Vaccine Sera and Monoclonal Antibodies. 2021; MedRxiv: doi: https://doi.org/10.1101/2021.12.07.21267432.

8. Cele S, Jackson L, Khoury DS, Khan K, et al. SARS-CoV-2 Omicron has extensive but incomplete escape of Pfizer BNT162b2 elicited neutralization and requires ACE2 for infection. medRxiv 2021.12.08.21267417; doi: https://doi.org/10.1101/2021.12.08.21267417

9. Sadoff J, Gray G, Vandebosch A, et al Safety and Efficacy of Single-Dose Ad26.COV2.S Vaccine against Covid-19 N Engl J Med 2021; 384:2187-2201 DOI: 10.1056/NEJMoa2101544

10. Bekker L, Garrett N, Goga AE, et al. Effectiveness of Ad26.COV2.S vaccine in health care workers in South Africa. the Lancet 2021: in press.

11. Johnson & Johnson Announces Real-World Evidence and Phase 3 Data Confirming Strong and Long-Lasting Protection of Single-Shot COVID-19 Vaccine in the U.S. https://www.jnj.com/:∼:text=The%20Phase%203%20ENSEMBLE%202,days%20after%20the%20first%20provided:&text=94%20percent%20protection%20against%20symptomatic,CI,%2058%-100%). (Accessed 24 December 2021)

12. Barouch DH, Stephenson K, Sadoff J et al. Durable Humoral and Cellular Immune Responses 8 Months after Ad26.COV2.S Vaccination. N Engl J Med 2021; 385:951-953 DOI: 10.1056/NEJMc2108829

